# X-linked multi-ancestry meta-analysis reveals tuberculosis susceptibility variants

**DOI:** 10.1101/2024.05.31.24308259

**Authors:** Haiko Schurz, Craig J Kinnear, Paul D van Helden, Gerard Tromp, Eileen G Hoal, International Tuberculosis Host Genetics Consortium, Marlo Möller

## Abstract

Globally, tuberculosis (TB) presents with a clear male bias that cannot be completely accounted for by environment, behaviour, socioeconomic factors, or the impact of sex hormones on the immune system. This suggests that genetic and biological differences, which may be mediated by the X chromosome, further influence the observed male sex bias. The X chromosome is heavily implicated in immune function and yet has largely been ignored in previous association studies. Here we report the first multi-ancestry X chromosome specific meta-analysis on TB susceptibility. We identified X-linked TB susceptibility variants using seven genotyping data sets and 20,255 individuals from diverse genetic ancestries. Sex-specific effects were also identified in polygenic heritability between males and females along with enhanced concordance in direction of genetic effects for males but not females. These sex-specific genetic effects were supported by a sex-stratified and combined meta-analysis conducted using the X chromosome specific XWAS software and a multi-ancestry analysis using the MR-MEGA software. Seven significant associations were identified. Two in the overall analysis (rs6610096, rs7888114) and a second for the female specific analysis (rs4465088) including all data sets. For the ancestry specific meta-analysis three significant associations were identified for males in the Asian cohorts (rs1726176, rs5939510, rs1726203) and one in females for the African cohort (rs2428212). Several genomic regions previously associated with TB susceptibility were reproduced in this study, along with strong ancestry-specific effects. These results support the hypothesis that the X chromosome and sex-specific effects could significantly impact the observed male bias in TB incidence rates globally.

## 1.1 Introduction

The X chromosome is estimated to encode approximately 1,500 of the 20,000 protein coding genes in the human genome (1) and has the highest density of regulatory miRNA (2). This means that the X chromosome codes for over 5% of proteins and 10% of miRNA respectively and even though many of these are involved in immune functions the X chromosome has generally been ignored in previous association studies (3). According to the Genome-Wide Association Study (GWAS) catalogue, 62,652 unique SNP-trait associations have been identified by 3,420 publications and of these associations only 385 SNPs were located on the X chromosome with 157 reaching genome-wide significance (p-value < 5*e*^−8^) (4). This indicates the extent to which the X chromosome has been ignored in GWAS even though it represents a significant portion of the genome. In the past this was due to the analysis complexities introduced to GWAS by the X chromosome due to male hemizygosity, but recently tools have been developed to analyse this chromosome specifically (5,6). Females have a more robust immune response against infections, which is partially attributable to the X chromosome and further influenced by the processes of X chromosome inactivation (XCI) and genes that escape silencing (7–11). Given the role of the X chromosome in sex-biased immune responses it should not be excluded from statistical analysis, especially for infectious diseases, which often presents with a male bias, and autoimmune diseases, which often presents with a female bias (11,12).

Tuberculosis (TB), caused by *Mycobacterium tuberculosis* (*M. tuberculosis*), presents with a strong male bias with the global reported incidence rate being nearly twice as high in males compared to females (13). This male bias has partially been attributed to socioeconomic and behavioural factors, access to healthcare and the effect of sex hormones (estrogen and testosterone) on the immune system (10,11,14,15). As these factors do not fully explain the sex bias, we hypothesise that X-linked genes and the process of XCI could further clarify the phenomenon. Seventeen published GWAS have investigated TB susceptibility (16–35), but only six of these included the X chromosome in their analysis (16,17,20,26,28,35) and only one of the studies focused specifically on the X chromosome (35). Our previous study, in a South African setting, revealed strong sex-specific effects on both the autosome and X chromosome, suggesting that sex-stratified quality control (QC) and association testing should be done when analysing the X chromosome and autosomes (35). These sex-stratified analyses are crucial as sex-specific effects of variants will be lost in a combined analysis, negating the opposite directions of effects between the sexes. As a result, vital information concerning TB susceptibility and sex bias is lost, highlighting the need for sex-stratified analysis.

Apart from ignoring the X chromosome, findings from previous studies have also validated poorly across populations. This is because ancestry has a major influence on TB susceptibility as shown in meta-analysis and admixture studies (36,37). Here, we report the first X chromosome-specific sex-stratified meta-analysis to identify TB susceptibility loci and elucidate its contribution to male sex bias, using GWAS data from European, Asian and African cohorts. Multi-ancestry meta-analyses have the advantage of giving an overview of both population-specific and global susceptibility variants and improving comprehension of the role of genetic ancestry and sex in TB susceptibility.

## 1.2 Methods

### 1.2.1 Study cohorts

Data for this meta-analysis was obtained from the International Tuberculosis Host Genetics Consortium (ITHGC) (17,25,28,29,38) and published TB GWAS (16,35). The ITHGC consists of 14 data sets, but eight cohorts had no X chromosome data or too few X-linked variants for imputation and were excluded from the analysis (39). In total two Asian, one European and four African cohorts, which included two five-way admixed cohorts from South African (SA) cohorts, were used for this meta-analysis (Table 1), with a total of 10791 cases, 12070 controls and a 1.85:1 male to female ratio. Approval for the study was obtained from the Health Research Ethics Committee of Stellenbosch University (project registration number S17/01/013 and 95/072).

**Table 1:** Genotyping platform and number of samples for each cohort prior to quality control and imputation.

| Country | Population | Cases | Controls | Females (%) | Platform | Source |
| --- | --- | --- | --- | --- | --- | --- |
| China (1) | Asian | 483 | 587 | 28.19 | Affymetrix SNP Array 6.0 | (29) |
| China (2) | Asian | 1290 | 1145 | 48.74 | Human OmniZhonghua-8 chips. | (29) |
| Russia | European | 5914 | 6022 | 29.32 | Affymetrix Genome-Wide Human SNP Array 6.0 | (17) |
| Gambia | African | 1316 | 1382 | 39.53 | Affymetrix GeneChip 500K | (28) |
| Ghana | African | 1359 | 1952 | 36.97 | Affymetrix SNP Array 6.0 | (25) |
| South Africa A | SA | 19 | 577 | 44.66 | Affymetrix 500k | (16) |
| South Africa M | SA | 410 | 405 | 55.78 | Illumina MEGA | (35) |

### 1.2.2 Quality control and Imputation

Quality control (QC) prior to imputation was performed as described previously (35), in order to obtain high quality genotypes for imputation. Briefly, the XWAS software (v3.0) and pipeline was used to implement a sex-stratified QC on all data, removing individuals without sex information and relatedness (up to 3^rd^ degree) (5,6). As the data sets had varying coverage of the X chromosome due to the different genotyping platforms used (Table 1), imputation was done to increase the number of overlapping variants for the meta-analysis. An in-house method employing Impute2 (40) and the full 1000 Genomes Phase 3 reference panel (41) was used for imputation as previously described (39,42), however data was not phased prior to imputation to increase imputation accuracy (40). Imputed variants were filtered at an info score of 0.45 and all non-binary variants were removed before converting the data to Plink format using GTOOL version 0.7.5 (43) at a genotype calling threshold of 0.7. Finally, the data underwent another sex-stratified QC procedure, using the same parameters as prior to imputation except that sex concordance and relatedness was not checked again. Overlapping variants between all data sets were extracted for the meta-analysis and aligned to the same reference allele where possible using the Plink v1.9 reference-allele command (44).

### 1.2.3 Association testing

A sex-stratified logistic regression test was performed for each data set using the XWAS (v3.0) software and incorporating the first five principle components as covariates for the ITHGC data and the four main ancestral components (Bantu-speaking African, KhoeSan, European and South Asian) for the SA. The SA was subject to sex-biased admixture events and as a result there are significant differences between the ancestral components of the autosome and X chromosome. Therefore, the ADMIXTURE software (version 1.3) was used to determine X chromosome specific ancestral components (using the --haploid flag), as described previously (35).

Since males are haploid for X-linked genes and females are diploid, females undergo unique biological processes to equalise the dosage of gene expression between the sexes by randomly silencing and inactivating one of the X-linked genes. This makes females functional mosaics for X-linked genes, and gene expression can be further altered by effects such as genes that escape silencing (leading to increased expression compared to males) or the inactivation can be skewed to favour a specific X chromosome (non-random inactivation) (11). To account for this variation and the sex-specific gene expression a statistical model to integrate different models of inactivation in the association testing model was used (35,45). In females each genomic location is modelled as either escaping inactivation, random inactivation or skewing towards the normal or deleterious allele and the model with the lowest log likelihood ratio is determined.

### 1.2.4 Meta-analysis

The meta-analysis was conducted by combining the association test results from each individual data set using the XWAS (v3.0) software (5,6). Association statistics for males and females were combined for the sex-stratified meta-analysis to determine sex-specific effects. In addition, all male and female results were combined into one meta-analysis to determine effects that are independent of sex. This was done in an ancestry-stratified and unstratified manner by testing all seven data sets together, or by grouping the two Asian or African cohorts (SA, Ghana, and Gambia). Since there is only one European data set a European-specific meta-analysis could not be performed. The Chi-squared based Q statistic was used to assess the heterogeneity between included studies. A fixed effect (FE) model was used when the p-value for heterogeneity was >0.1 otherwise a random effects (RE) model was used to calculate pooled odds ratio (OR) and p-values (46). Finally, due to the different genetic ancestry of the input data sets (Table 1 and 2) the MR-MEGA multi-ancestry meta-regression software was implemented as described previously (39,47), using genomic control correction (for individual input data sets with lambda inflation values >1.05) and including the first two principal components (calculated from the genome-wide allele frequency differences) as covariables in the regression. The genome-wide significance threshold for the meta-analysis was set to 5*e*^−8^ (48).

### 1.2.5 Polygenic heritability

To determine whether the level of genetic contribution to TB susceptibility differs between males and females for the autosomal chromosome only and for the autosomal chromosome including the X chromosome, polygenic heritability on the individual data sets were estimated (Table 1 and 2). Polygenic heritability estimates, for the combined and sex-stratified data sets, were calculated using GCTA (v1.93.2), a genomic risk prediction tool (49). The genetic relationship matrix was calculated for each autosomal chromosome and the X chromosome (combined and sex-stratified data) prior to being merged and filtered by removing cryptic relatedness (--grm-cutoff 0.025) and assuming that the causal loci have similar distribution of allele frequencies as the genotyped SNPs (--grm-adj 0). Principal components were then calculated (--pca 20) to include as covariates prior to estimating heritability. The average heritability estimate was calculated by taking the mean and calculating the standard deviation of all estimates. The heritability estimates of the autosomal chromosomes and the autosomal plus X chromosome were compared between males, females, and combined data sets to determine if there are any significant sex-specific effects on heritability.

### 1.2.6 Concordance in direction of effects

To determine the degree to which direction of effects of SNPs on the X chromosome is shared between the ancestry-specific XWAS meta-analysis we followed the methodology originally developed by Mahajan et.al. (50), which we implemented previously (39). Since the ITHGC data only had one European data set with an X chromosome available we could not do a European ancestry meta-analysis, so for this analysis we used the Russian data set to present the European component. For each ancestry specific meta-analysis (and Russian data association statistics) independent variants were identified (separated by at least 500kb) at different p-value thresholds (0.001 < P ≤ 0.01; 0.01 < P ≤ 0.5; and 0.5 < P ≤ 1). The identified independent SNPs were extracted from all other data sets and significant excess in concordance of direction of effect was tested using a one-sided binomial test with an expected frequency of 50%. The analysis was done for the combined and sex-stratified data, using each of the ancestral analysis as reference to identify independent SNPs.

## 1.3 Results

### 1.3.1 Cohort summary

Summary statistics of all the data sets following imputation and QC are shown in Table 2. In total 20,255 individuals passed QC (9468 cases and 10787 controls) of which 35% were female. Between the different data sets 69,983 variants overlapped and aligned to the same reference allele.

**Table 2:**
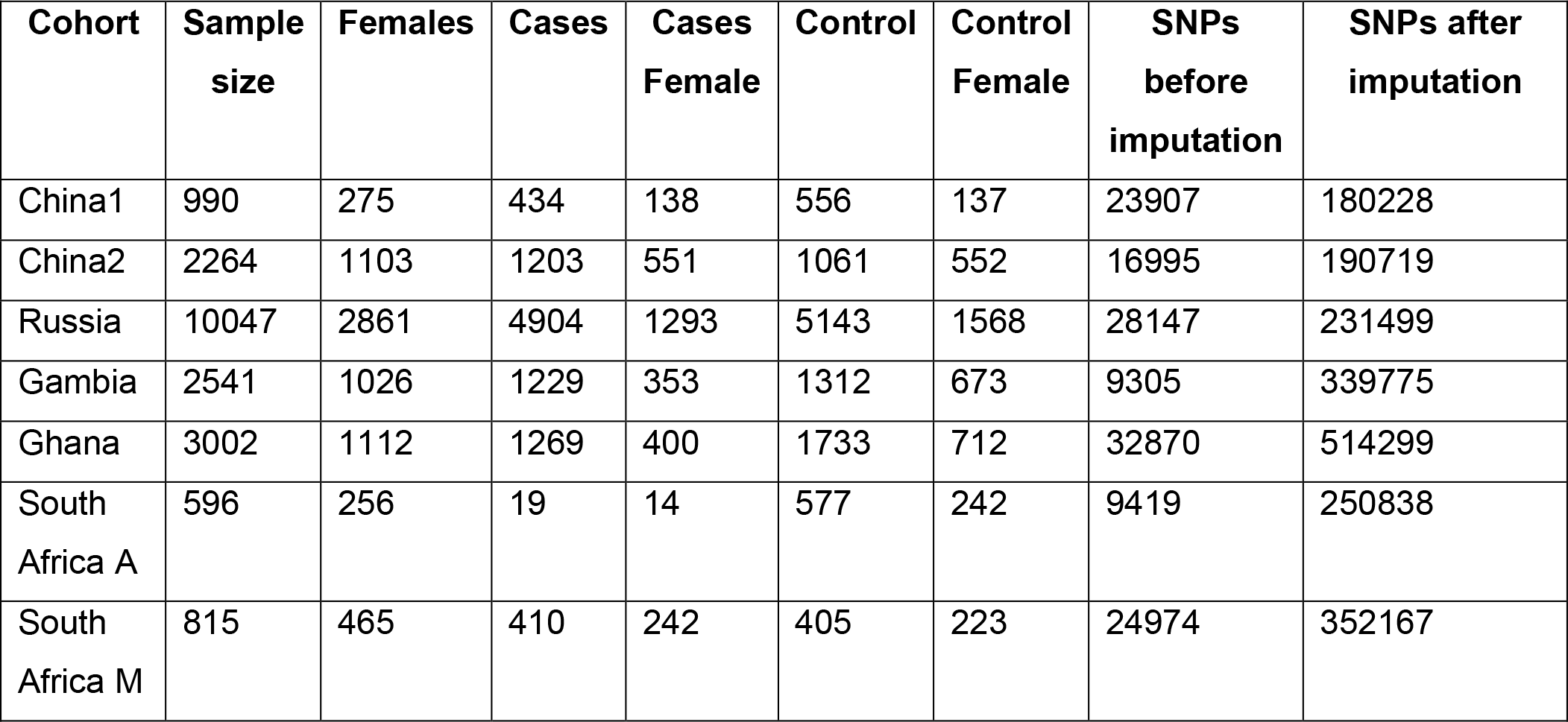
Cohort summary and number of X chromosome variants post Imputation and QC.

| Cohort | Sample size | Females | Cases | Cases Female | Control | Control Female | SNPs before imputation | SNPs after imputation |
| --- | --- | --- | --- | --- | --- | --- | --- | --- |
| China1 | 990 | 275 | 434 | 138 | 556 | 137 | 23907 | 180228 |
| China2 | 2264 | 1103 | 1203 | 551 | 1061 | 552 | 16995 | 190719 |
| Russia | 10047 | 2861 | 4904 | 1293 | 5143 | 1568 | 28147 | 231499 |
| Gambia | 2541 | 1026 | 1229 | 353 | 1312 | 673 | 9305 | 339775 |
| Ghana | 3002 | 1112 | 1269 | 400 | 1733 | 712 | 32870 | 514299 |
| South Africa A | 596 | 256 | 19 | 14 | 577 | 242 | 9419 | 250838 |
| South Africa M | 815 | 465 | 410 | 242 | 405 | 223 | 24974 | 352167 |

### 1.3.2 Individual data set association results

Five genome-wide significant variants were identified for the individual data sets (Table 3). Manhattan and quantile-quantile (QQ) plots for these results are given in supplementary Figures S1 and S2. One SNP (rs4465088) is located downstream of the actin related protein T1 gene (*ACTRT1)* in the Ghanaian females (OR = 4.73, p-value = 4.53*e*^−18^) and in the combined analysis (p-value = 1.60*e*^−16^) (Fisher’s method for combining p-values). *ACTRT1* encodes for a protein related to cytoskeletal beta-actin and while it is also involved in spermatid formation this gene has been shown to have tumour suppressive properties. The tumour suppressive properties were identified in basal cell carcinomas which result from the aberrant activation of the Hedgehog signalling pathway (51). While these genes have not been previously implicated in TB susceptibility, the sonic hedgehog pathway (homolog of the hedgehog pathway) is implicated in mycobacterial immune evasion mediated through exploitation of regulatory T-cells (52).

**Table 3:** Results for X chromosome association testing of individual data sets using the XWAS software for the sex-stratified analysis and combined p-value using the Fisher’s method.

| SNP | Gene (locus) | Position | A1 | Male |  |  | Female |  |  | P comb | Cohort |
| --- | --- | --- | --- | --- | --- | --- | --- | --- | --- | --- | --- |
|  |  |  |  | OR* | P-value | 95% CI* | OR | P-value | 95%CI |  |  |
| rs4465088 | *ACTRT1* (Xq25) | 3'UTR | G | 0.94 | $8.58e^{-1}$ | 0.46-1.91 | 4.73 | $4.53e^{-18}$ | 3.42-6.55 | $1.60e^{-16}$ | Ghana |
| rs6610096 | *PAGE4* (Xp11.23) | 5'UTR | A | 1.53 | $9.95e^{-14}$ | 1.37-1.72 | 1.70 | $8.95e^{-16}$ | 1.49-1.94 | $5.84e^{-27}$ | Russia |
| rs7888114 | *FRMPD4* (Xp22.2) | Intron | C | 19.29 | $9.72e^{-9}$ | 7.05-52.8 | 3.14 | $1.18e^{-3}$ | 1.57-6.31 | $3.00e^{-10}$ | Russia |
| rs2563800 | *LOC105373293*<br>(Xq21.31) | Intron | A | 1.57 | $1.08e^{-8}$ | 1.35-1.83 | 1.04 | 0.659 | 0.87-1.25 | $1.41e^{-7}$ | Russia |
| rs5975736 | *BRS3* (Xq26.3) | 3'UTR | A | 2.49 | $6.1e^{-7}$ | 1.74-3.56 | 2.29 | $3.43e^{-5}$ | 3.52-49.2 | $5.36e^{-10}$ | Russia |

\*OR: Odds ratio; 95% CI: 95% confidence interval

Four significant variants were identified in the Russian cohort, three of which were significant in males (rs6610096, rs7888114, rs2563800), one in females (rs6610096) and two for the combined test statistic (rs6610096, rs5975736). The rs6610096 variant, located in the prostate associated gene 4 (*PAGE4*) was significant in male, females, and the combined analysis. *PAGE4* is a member of the cancer/testis X antigens and while its function is poorly understood it is a DNA binding protein and silencing of *PAGE4 in vitro* caused cell death by apoptosis, indicating that *PAGE4* has an anti-apoptotic function (53). Expression of *PAGE4* is also inversely correlated with cancer progression and dysregulation of *PAGE4* modulates androgen receptor signalling and promotes progression to advanced prostate cancer (54). The rs2563800 variant, significantly associated in males and the combined analysis, is located in a non-coding RNA (ncRNA) *LOC105373293*, with no known function. *FRMPD4,* a gene previously implicated in schizophrenia, was only significantly associated in males (rs7888114), while rs5975736, located in the Bombesin Receptor Subtype 3 (*BRS3)* gene was associated in the combined data set and had the same direction of effect between the sexes (55). *BRS3* is involved in energy homeostasis and upregulates glucose-stimulated insulin secretion in human pancreatic islet cells and could have a potential role in treatment of obesity and diabetes mellitus (56,57).

In addition to sex-stratified analysis we also modelled patterns of XCI in females to incorporate in the association testing. X chromosome inactivation is the process through which one X chromosome is randomly inactivated in females to equalise the dosage of gene expression between males and females (11). However, the process of XCI can suffer from effects such as genes escaping inactivation or skewed inactivation where on X chromosome is disproportionately silenced, which can have significant effects on gene expression, the immune response and disease susceptibility between the sexes (11). Using this method we identified one genome-wide association in the Ghanaian cohort, rs4465088, which is the same association identified by the sex-stratified analysis of this data set. The analysis suggests that this variant is most likely experiencing escape from inactivation, which suggests that it is overexpressed in females compared to males. This may explain why this variant was associated in females only in the sex-stratified analysis (Table 4). While not reaching genome-wide significance the next lowest p-values were all unique associations not identified as top hits in any other analysis done in this paper including rs266786, located in the HECT, UBA and WWE domain containing E3 ubiquitin protein ligase 1 (*HUWE1)* gene. While the *HUWE1* gene has not been implicated in TB susceptibility it does promote host defence against bacterial infection by mediating inflammasome activation and could be a viable candidate gene for future TB research (58).

**Table 4:** Results for the association testing for individual data sets, incorporating X chromosome inactivation modelling into the analysis.

| SNP | Position | Gene | Model | OR | 95% CI | P-value | Cohort |
| --- | --- | --- | --- | --- | --- | --- | --- |
| rs4465088 | 3'UTR | *ACTRT1* (Xq25) | Escape of XCI | 0.28 | 0.21-0.38 | $1.57e^{-15}$ | Ghana |
| rs266786 | Intron | *HUWE1* (Xp11.22) | Skewed XCI to risk allele | 1.55 | 1.31-1.84 | $3.07e^{-07}$ | Ghana |
| rs5935414 | 3'UTR | *MRPL35P4* (Xp22.2) | Skewed XCI to risk allele | 2.06 | 1.57-2.75 | $3.32e^{-07}$ | Gambia |
| rs183350242 | 3'UTR | *IRS4* (Xq22.3) | Skewed XCI to risk allele | 0.16 | 0.08-0.34 | $4.60e^{-07}$ | China1 |
| rs5963484 | 3'UTR | *UBTFL11* (Xp11.4) | Random XCI | 0.86 | 0.81-0.91 | $8.96e^{-07}$ | Russia |

### 1.3.3 Polygenic heritability

Twin studies have estimated that the narrow sense heritability of susceptibility to TB can be up to 80%, but very few modern estimates have been done and none have focused on sex-stratified heritability estimates (26,39,59–61). In our a previous meta-analysis we estimated the polygenic heritability of TB susceptibility on the autosomal chromosomes of the ITHGC data, which ranged from 5-36% (39). Using the same methods and WHO prevalence rates for each country (Table S1) we expand on the polygenic heritability analysis here for all data sets where we have the X chromosome data available. Heritability estimates were obtained for the autosomal chromosomes and for the autosomal chromosomes including the X chromosome and both analyses were performed on a sex-stratified and combined data set. Including the X chromosome into the analysis did not significantly change the heritability estimates (Table S1), however for both the autosomal and autosome plus X chromosome analysis the heritability estimates were significantly different between males and females (Table S1, Figure S3).

Polygenic heritability for males was lower compared to females and while environmental factors between studies from different geographical locations contributes to the variation in heritability estimates between the input data, the same argument cannot be made for the sex-stratified analysis. The influence of environmental factors is minimized because males and females for the individual data sets were sampled from the same region. This suggests that genetic components are not the main driving factors for the global sex bias in TB incidence rates, and that non-genetic factors such as behavioural differences have a larger impact on the sex bias. Identifying and including these non-genetic factors in statistical analysis could help identify which factors have the biggest impact and highlight any underline genetic effects. Furthermore, future in depth polygenic heritability of susceptibility to TB should focus on sex-specific analysis as a combined analysis masks the different heritability estimates between males and females.

### 1.3.4 Meta-analysis results

Both a genome-wide significance threshold (5*e*^−8^) and a Bonferroni threshold (7.14*e*^−7^) were implemented to correct for the number of variants tested in the meta-analysis (0.05/69983). Since TB is a complex disease and variants are unlikely to have large effect sizes, lowering the significance threshold can help to reduce the loss of valuable information in the form of false negative results. In total four variants reached significance in the sex-stratified population specific analysis, while no significant associations were identified in the combined (male and female) and sex-stratified analysis including all data sets (Table 5 and 6). The QQ and Manhattan plots for these association tests are given in supplementary Figures S4-S6. The most significantly associated variant for the non-sex-stratified analysis including all data sets was rs79720685 (OR = 0.83, p-value = 3.06*e*^−5^) located in the interleukin 1 receptor accessory protein like 1 (*IL1RAPL1*) gene (Table 5). Variants in this gene have been associated with cardiovascular disease (62), autism (63) and presented with a male sex bias in a previous XWAS of asthma susceptibility in children (3). Variants in *IL1RAPL1* downregulate interleukin (IL) 13 which has a negative impact on the IL-1R pathway, a potential regulator of inflammation and a critical component of the host innate immune response against infections (64). The impact of this gene on TB susceptibility is unclear but given its role in the immune response it may contribute to the disease. The top hit for the combined meta-analysis in the Chinese cohorts was related to spermatogenesis and thus not informative in the context of TB susceptibility (65). For the African cohorts the combined analysis revealed the variant with the lowest p-value to be in the actin remodelling regulator *NHS* gene, previously associated with Nanca-Horan Syndrome (a congenital cataract disease), dental abnormalities, brachymetacarpia (an abnormal shortness of the metacarpal bones) and mental retardation (66–69).

**Table 5:** Overall and ancestry-specific meta-analysis results for the combined analysis using the X chromosome specific XWAS software.

| SNP | Gene (locus) | Position | A1 | N | P-value | OR* | Q | Model | cohort |
| --- | --- | --- | --- | --- | --- | --- | --- | --- | --- |
| rs79720685 | *IL1RAPL1*<br>(Xp21.3-21.2) | Intron | T | 14 | $3.06e^{-5}$ | 0.83 | 0.37 | Fixed | All |
| rs58085560 | *SPANXN2*<br>(Xq27.3) | 3'UTR | C | 4 | $2.85e^{-5}$ | 1.32 | 0.86 | Fixed | China |
| rs5909376 | *NHS*<br>(Xp22.2-22.13) | Intron | T | 8 | $4.11e^{-5}$ | 1.22 | 0.63 | Fixed | African SA |

\*OR: Odds ratio

For the sex-stratified meta-analysis the variants with the lowest p-values in the analysis including all cohorts for males was rs753468 (OR_M = 0.84, p-value = 3.21*e*^−5^), located in *ATRX* and for females rs7053675 (OR_F = 0.83, p-value = 5.56*e*^−6^) located in the *PTCHD1-AS* gene (Table 6). The chromatin remodeller *ATRX* plays a role in supressing deleterious DNA secondary structures that form a transcribed telomeric repeat, and loss of function of the *ATRX* gene can increase DNA damage and stall replication and homology-directed repair (70). This suggests that *ATRX* is involved in essential biological processes and it has been previously implicated in intellectual disability and osteosarcoma (71). Disruptions of the *PTCHD1-AS* gene are prevalent in ∼1% of Autism spectrum disorders and intellectual disabilities (72). For the Chinese cohorts the sex-stratified analysis revealed 3 genome-wide significant associations (rs1726176, rs5939510, rs1726203) in the long intergenic non-protein coding RNA 1546 (*LINCO1546*) in males, while no significant associations were identified in females. The close proximity of these variants in *LINC01546* suggest LD between the variants and it is unclear if any of the variants influence TB susceptibility as no functional information is available for the *LINC01546* locus. Finally, in the African cohorts one variant, in the *UPF3B* gene, reached significance in females after Bonferroni correction (rs2428212, OR_F = 2.03, p-value = 4.72*e*^−7^) but not in males. *UPF3B* is a regulator of nonsense mediated mRNA decay (NMD) and rapidly breaks down aberrant mRNA with a premature termination codon (PTC). *UPF3B* is involved in a central step in RNA surveillance by regulating crosstalk between the NMD pathway and the PTC-bound ribosome complex (73). Mutations in the *UPF3B* gene disrupt the NMD pathway, which is critical for neuronal development and can cause various forms of intellectual disability (74–76). While this gene has not previously been implicated in TB susceptibility, it could influence disease by altering RNA regulation linked to host defence against TB.

**Table 6:** Overall and ancestry-specific meta-analysis results for the sex-stratified analysis using the X chromosome specific XWAS software.

| SNP | Gene (locus) | Position | A1 | N | Male |  | Female |  | Q (M/F) | Model | Cohort |
| --- | --- | --- | --- | --- | --- | --- | --- | --- | --- | --- | --- |
|  |  |  |  |  | OR* | P | OR | P |  |  |  |
| rs753468 | *ATRX* (Xq21.1) | Intron | C | 7 | 0.84 | $3.21e^{-5}$ | 1.06 | $1.62e^{-1}$ | 0.42/0.66 | Fixed | All |
| rs7053675 | *PTCHD1-AS* (Xp22.11) | Intron | A | 7 | 1.05 | $2.11e^{-1}$ | 0.83 | $5.56e^{-6}$ | 0.15/0.01 | Fixed | All |
| rs1726176 | *LINC01546* (Xp22.33) | 3'UTR | A | 7 | 1.82 | $4.20e^{-8}$ | 0.97 | $7.19e^{-1}$ | 0.80/0.75 | Fixed | China |
| rs5939510 | *LINC01546* (Xp22.33) | 3'UTR | A | 7 | 1.82 | $4.89e^{-8}$ | 0.98 | $7.50e^{-1}$ | 0.79/0.74 | Fixed | China |
| rs1726203 | *LINC01546* (Xp22.33) | 3'UTR | G | 7 | 1.81 | $6.37e^{-8}$ | 0.97 | $6.92e^{-1}$ | 0.86/0.69 | Fixed | China |
| rs2428212 | *UPF3B* (Xq24) | Intron | C | 7 | 0.76 | $2.28e^{-1}$ | 2.03 | $4.72e^{-7}$ | 0.95/0.15 | Fixed | African SA |

\*OR: Odds ratio

Due to the shared polygenic heritability across the data sets, we used MR-MEGA (Meta-Regression of Multi-Ethnic Genetic Association, v0.20), a meta-analysis tool that maximizes power and enhances fine-mapping when combining data across different ancestries. Despite MR-MEGA not having X chromosome specific methodologies two genome-wide significant associations (also identified in the analysis of individual data sets, Table 3) were identified for the combined (rs6610096) and female specific analysis (rs4465088, Table 7, Figure S7). In comparison the X chromosome specific analysis did not identify any significant associations when all data sets were included, which highlights the impact that genetic ancestry has on the results. The impact of ancestry is made clear when observing the p-value for ancestral heterogeneity in the MR-MEGA analysis, which is generally much lower than the p-value for residual heterogeneity. Future methods should thus develop X-chromosomal analysis that can accurately account for and maximise power across genetic ancestries. The MR-MEGA analysis also identified one association (rs7888114, Table 7), for the Bonferroni corrected significance threshold, located in the FERM and PDZ domain containing 4 gene (*FRMPD4*) in the combined analysis. While the potential impact of this gene on TB susceptibility is unclear a previous analysis by our group identified this gene in a SNP interaction analysis done on the SA data (35).

**Table 7:** Results for the combined and sex-specific analysis including all cohorts in for the MR-MEGA meta-analysis to adjust for effects of genetic ancestry.

| SNP | Gene | Position | A1 | N | P-value association | P-value ancestry het | P-value residual het | InBF | Analysis |
| --- | --- | --- | --- | --- | --- | --- | --- | --- | --- |
| rs6610096 | *PAGE4*<br>(Xp11.23) | 5'UTR | A | 6 | 1.90e<sup>-17</sup> | 2.69e<sup>-04</sup> | 0.86 | 48.71 | Combined |
| rs7888114 | *FRMPD4*<br>(Xp22.2) | Intron | C | 10 | 1.95e<sup>-07</sup> | 8.44e<sup>-08</sup> | 0.03 | 13.58 | Combined |
| rs5939515 | *MXRA5*<br>(Xp22.33) | 5'UTR | A | 6 | 1.05e<sup>-06</sup> | 3.53e<sup>-07</sup> | 0.99 | 12.46 | Male |
| rs5939197 | *MXRA5*<br>(Xp22.33) | 5'UTR | A | 6 | 1.64e<sup>-06</sup> | 6.90e<sup>-07</sup> | 0.94 | 12.01 | Male |
| rs4465088 | *ACTRT1*<br>(Xq25) | 3'UTR | T | 7 | 1.97e<sup>-08</sup> | 2.71e<sup>-07</sup> | 5.52e<sup>-10</sup> | 16.02 | Female |

### 1.3.5 Concordance in direction of effects

In a previous publication by the ITHGC concordance in direction of effects was assessed on the autosomal chromosomes and no significant effects were identified (Supplementary Table S2-S4). Here we repeated the analysis on the X chromosome for both the combined and sex-stratified data sets. No significant effects were identified for the combined and female-specific analysis. For the male analysis significant concordance was identified between the Asian and African data sets for variants with p-values smaller than 0.001 (using the Asian data set as reference, p-value = 0.007) and between the European and African data sets for variants with p-values between 0.001 and 0.01 (using the European data set as reference, p-value 0.049). Furthermore, while the p-values for the combined and female specific analysis were similar but overall, the p-values for males were lower, suggesting stronger concordance in direction of effects on the X chromosome for males.

## 1.4 Discussion

Here we report the first X chromosome specific meta-analysis to investigate human genetic susceptibility to TB and the observed male bias. Two significant variants were identified across all cohorts for the combined and females specific MR-MEGA analysis (Table 7), while no susceptibility variants reached significance in the X chromosome specific meta-analysis including all cohorts. When stratified by sex and population group four variants were identified using the X chromosome specific analysis, three variants for males in the Chinese cohorts and one variant in females in the African cohorts (Table 6 and 7). Analysis of the individual data also revealed five significant associations in the Ghanaian and Russian data sets (Table 3 and 4).

While most of the genes identified in this study have not been previously implicated in TB susceptibility a few could potentially be involved in mechanisms associated with the disease. *ACTR1, IL1RAPL1, ATRX, UPF3B* and *HUWE1* are involved in cellular functions that could be linked to host defence against TB. *ACTR1* and *IL1RAPL1* are involved in immune pathways via the sonic hedgehog signalling pathway and IL-1R pathway respectively, and mutations can affect T-cell regulation (52) and host immune response to infection (64), both of which are involved in host defence against TB. *ATRX* and *UPF3B* are involved in essential biological processes by monitoring and controlling aberrant RNA that could negatively impact transcription and RNA regulation (70,73).

*HUWE1* is involved in inflammasome activation and important for Caspase-1 maturation and IL-1β release and previous studies have shown that both mice and human cell lines experienced increased bacterial burden and decreased caspase-1 activation and IL-1β release (58). During infection *M. tuberculosis* can directly influence caspase-1/IL-1β inflammasome activation, making HUWE1 a viable candidate for future research in TB susceptibility (77,78). The role of these genes in immune function and RNA regulation suggests that they could impact TB susceptibility, but further investigation is required to elucidate the functional mechanisms underlying our statistical findings.

Previous TB susceptibility studies investigating X-linked genes have identified several *TLR8* variants, but these associations were not replicated in this study (79–84). Possible reasons for this could be the impact of population-specific effects and more stringent significance thresholds. However, while the exact variants and genes previously identified with TB susceptibility on the X chromosome were not replicated, the genomic regions where these genes are located did replicate. In a linkage study the genomic region Xq, specifically Xq26, was associated with TB susceptibility in an African cohort (85). Indirect evidence of TB susceptibility loci on the X chromosome has also been provided from studies in Mendelian susceptibility to Mycobacterial Diseases (MSMD) where two X-linked regions, Xp11.4-Xp21.2 and Xq25-26.3 have been associated with MSMD (86,87). All these genomic regions were validated in this study, as well as the genomic locus where *TLR8* is situated (Xp22). We identified significant associations at Xp11.22-Xp11.4, Xp21-Xp22.33, Xq21 and Xq24-Xq27.3, which overlap with previously associated genomic regions. This suggests that these genomic regions are implicated in TB susceptibility, but further research and fine-mapping is required to elucidate which genes and variants in this region contribute to the phenotype.

Many X-linked genes have not been fully characterised, and their functions are still unclear; a recurring theme in our study. As a result, the impact on TB susceptibility cannot be elucidated using bioinformatic analysis alone and functional analysis of the genes is required. This is a major limitation in XWAS as many significant associations cannot be fully elucidated without functional verification, due to the lack of information about X-linked genes and their involvement in biological mechanisms. The impact of a variant on gene function can be tested by introducing the variant into an appropriate cell line (in vitro) or animal model (in vivo) using genomic editing methods such as CRISPR (88,89). Infection studies using these edited cell lines or animals can be done to determine the function of the variant. Future studies could also include the Y chromosome in sex-specific analysis. While previous studies suggested that the Y chromosome does not have immune-related genes that could influence the sex bias, recent studies have significantly improved coverage of the Y chromosome, adding over 30 million base pairs and 42 protein coding genes (90). This improved reference sequence, along with additional clinical variants and functional genomics suggests that the impact of the Y chromosome on sex-specific effects should be revisited in future studies.

A second conclusion that can be drawn from these results is that there are strong population-specific effects influencing TB susceptibility. The fact that no significant associations were identified when all studies were included in the analysis, unless the genetic ancestry was accounted for in the MR-MEGA analysis, supports this hypothesis. Previous studies in admixed populations have also shown increased susceptibility for some ancestral components over others (36). While the impact of genetic ancestry cannot be ignored and population stratified analysis are vital, the fact that all populations show a degree of polygenic heritability for susceptibility to TB suggests that both global and population specific susceptibility must be investigated to elucidate the full complexity of TB disease. Identifying globally associated susceptibility variants can help reduce the global TB burden, and in combination with precision medicine using tailored and targeted treatments for specific population groups and sexes (91), especially in high burden settings can have a profound impact on the fight against TB.

We have shown in a previous XWAS in the SA population (35) that strong sex-specific effects exist, and this is mirrored in this study. When comparing the OR of males and females (Table 3 and 5) it is clear that many sex-specific variants have opposite directions of effect or a negligible effect in one of the sexes. We found 6 significant associations in males but only 3 in females and one in the combined analysis (when accounting for genetic ancestry), making a strong case for sex-specific effects. Sex-specific effects are further supported by the fact that there are significant differences in polygenic heritability for susceptibility to TB between the sexes and the overall higher level of concordance in direction of genetic effects on the X chromosome for males compared to females. A third implication, that has been discussed in detail elsewhere, is that careful phenotyping of cases and controls are necessary, both when studying *M.tuberculosis* infection and active TB (92).

We suggest that during the planning of TB susceptibility studies, power should be determined based on the sample size of one sex, to maintain enough statistical power for sex-stratified analysis. Furthermore, care should be taken during sample selection to minimise population specific effects and subsequent larger and more powerful multi-ancestry meta-analysis will need to be performed to identify global susceptibility variants. Finally, a multi-ancestry, X chromosome specific meta-analysis tool should be developed to identify population specific, sex-specific, and global susceptibility variants to elucidate some of the complexity of TB pathogenesis and eventually allow for tailored or even preventative treatment.

## Data Availability

Summary statistics of all analysis will be made available on the Dryad online database, and the summary statistics and original data of the individual data sets can be requested through the corresponding authors (Table 1 and 2).

## Acknowledgement

We would like to acknowledge and thank the study participants for their contribution and participation. This research was partially funded by the South African government through the South African Medical Research Council. The content is solely the responsibility of the authors and does not necessarily represent the official views of the South African Medical Research Council. This work was also supported by the National Research Foundation of South Africa (grant number 93460) to EH. This work was also supported by South African Tuberculosis Bioinformatics Initiative (SATBBI), a Strategic Health Innovation Partnership grant from the South African Medical Research Council and Department of Science and Technology to GT. We would also like to thank and acknowledge Dr Vivek Naranbhai and the International Tuberculosis Host Genetic consortium for allowing us access to their TB GWAS data.

The authors report no conflict of interest.

## 1.5 Ethics and data sharing

A research collaboration agreement was signed by all contributors. Ethics approval for the meta-analysis presented here was granted by the Health Research Ethics Committee of Stellenbosch University (project registration number S17/01/013). In addition, all institutions involved in the ITHGC have ethics approval for their respective studies:

China 1 and 2: The study protocol was approved by the Ethics Committee of the Beijing Chest Hospital, the 309 Hospital of the PLA, Shijiazhuang Fifth Hospital, the China PLA General Hospital, the Tongliao TB institute and the Center for Diseases Control and Prevention in Jalainuoer.

Russia: Blood samples from all participants were collected and studied with written informed consent according to the Declaration of Helsinki and with approvals from the local ethics committees in Russia (St. Petersburg and Samara) and the UK (Human Biological Resource Ethics Committee of the University of Cambridge and the National Research Ethics Service, Cambridgeshire 1 REC, 10/H0304/71).

Gambia: Ethics approval was granted by the Medical Research Council (MRC) and the Gambian government joint ethical committee.

Ghana: Ethics approval was granted by the Committee on Human Research, Publications and Ethics, School of Medical Sciences, Kwame Nkrumah University of Science and Technology, Kumasi, Ghana, and the Ethics Committee of the Ghana Health Service, Accra, Ghana.

SA A and SA M: Ethics approval was granted by the Health Research Ethics Committee of Stellenbosch University (project registration numbers S17/01/013, NO6/07/132 and 95/072).

## Supplementary material

**Figure S1:**
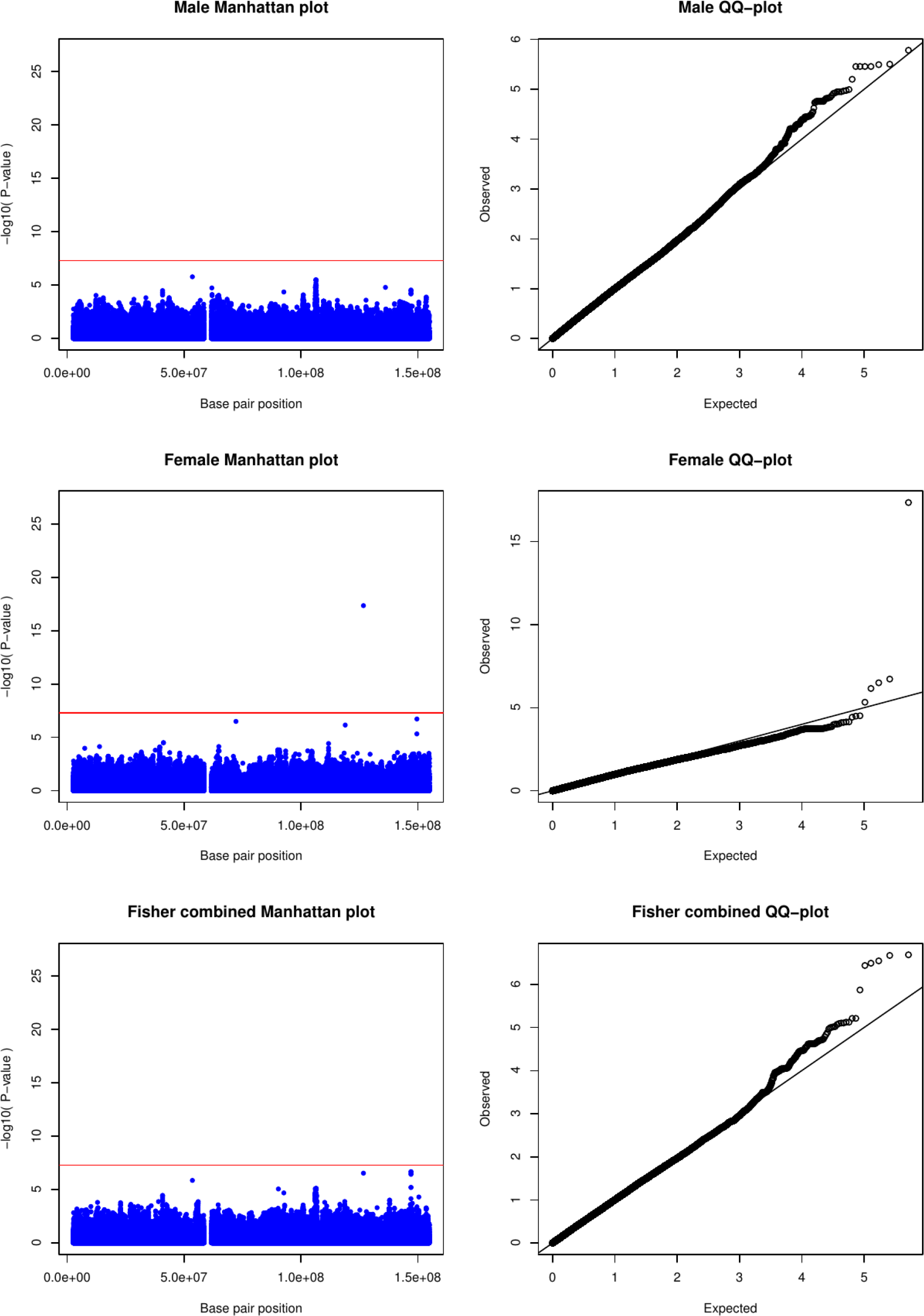
Manhattan and QQ-plot for X-linked SNP association testing of the Ghanaian cohort using the X chromosome specific XWAS software.

**Figure S2:**
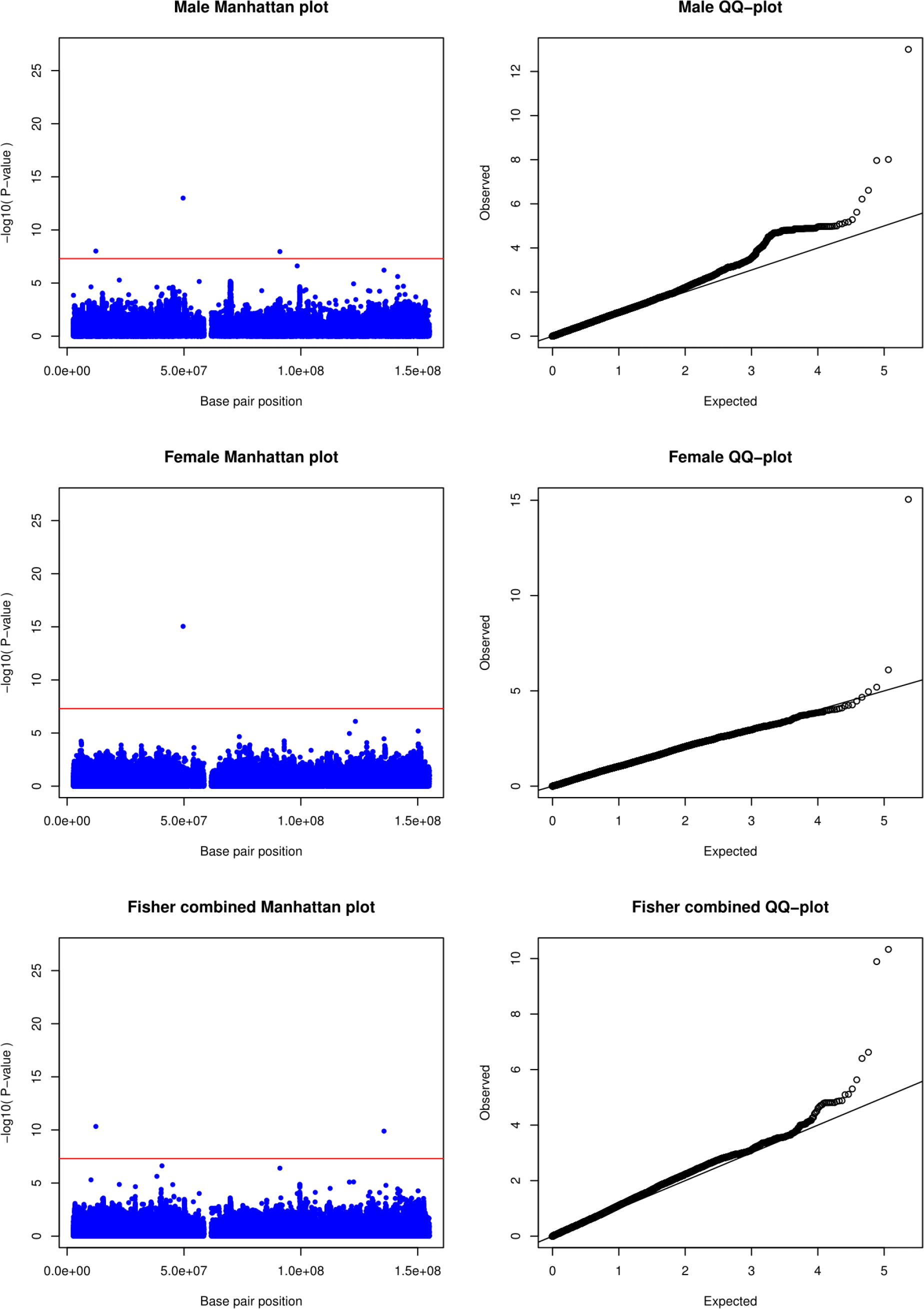
Manhattan and QQ-plot for X-linked SNP association testing of the Russian cohort using the X chromosome specific XWAS software.

**Figure S3:**
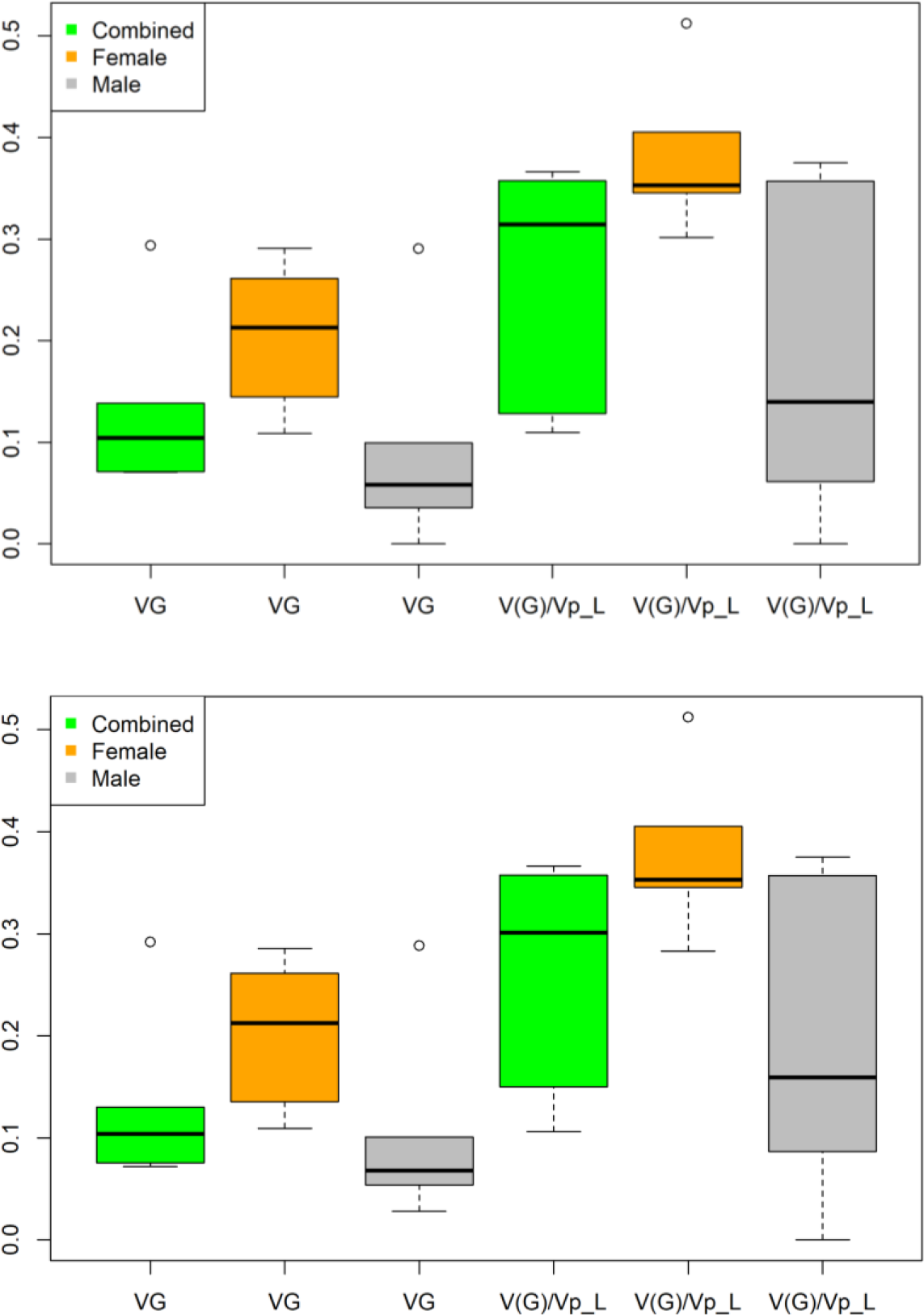
Distribution of genetic variance (VG) and Ratio of genetic variance to phenotypic variance (V(G)/Vp_L) across all data sets for male’s, females and combined data sets on autosomal chromosomes only [top] and autosomal chromosomes and the X chromosome [bottom].

**Figure S4:**
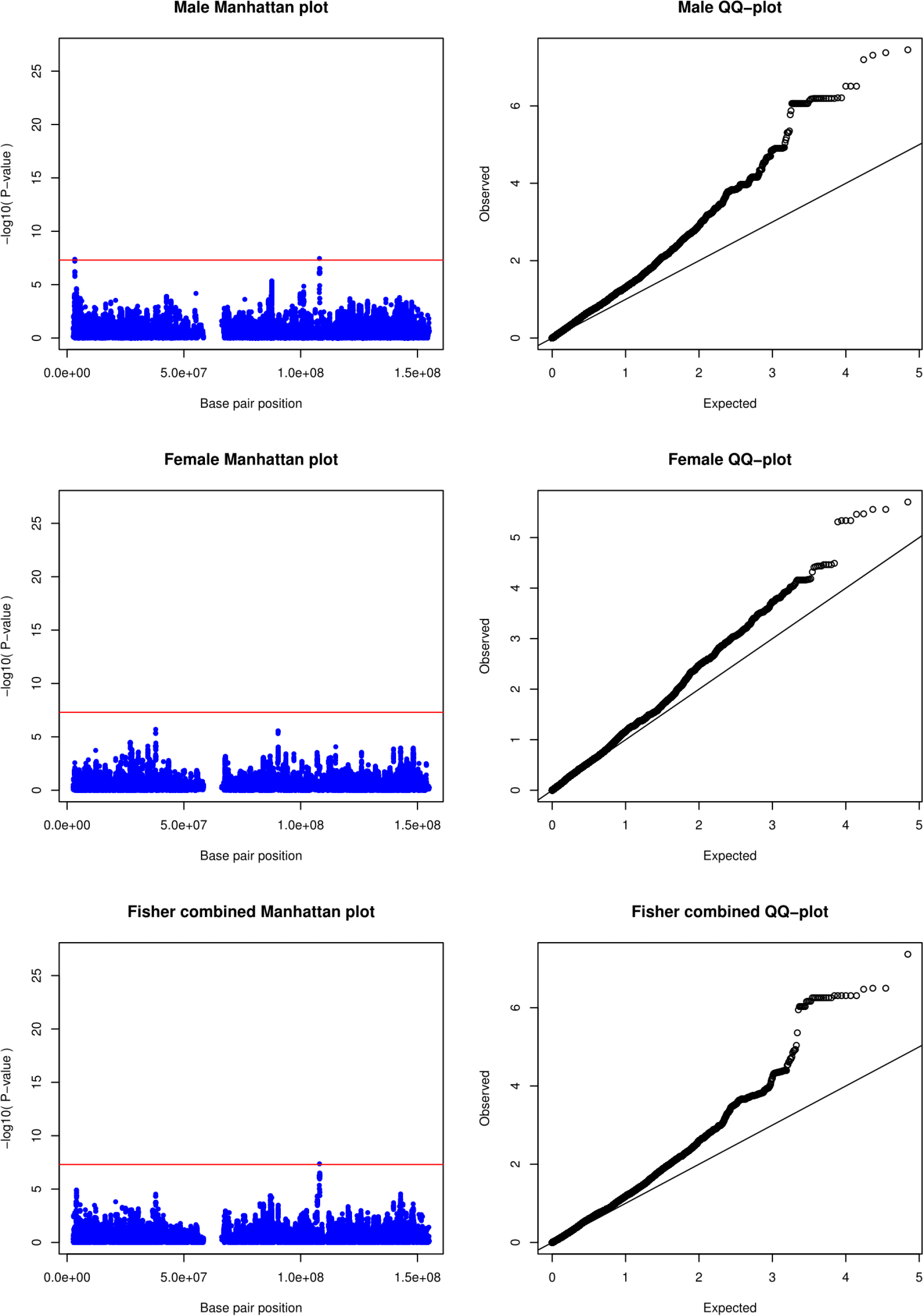
Manhattan and QQ-plot for the X-linked meta-analysis of the Chinese cohorts using the X chromosome specific XWAS software.

**Figure S5:**
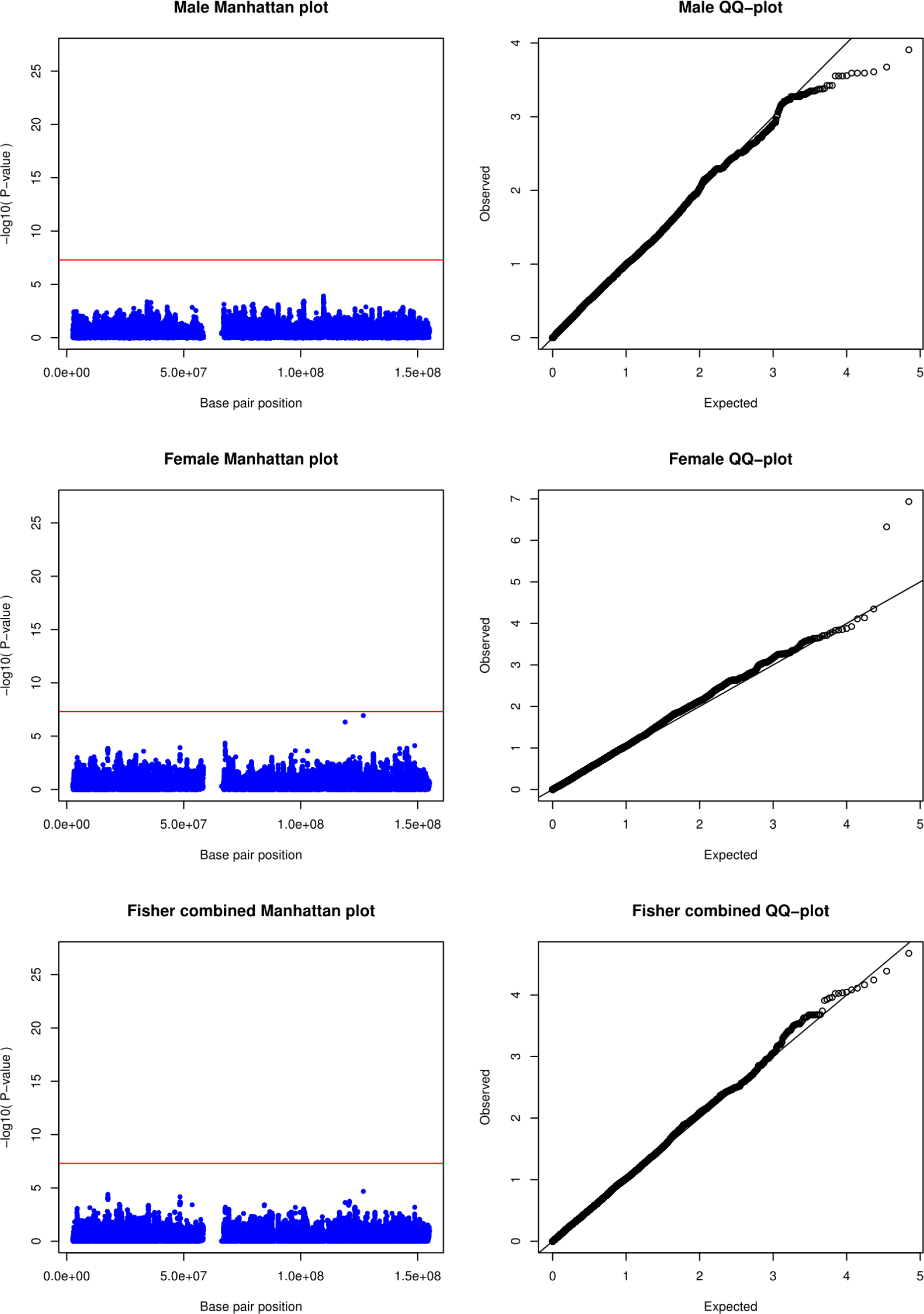
Manhattan and QQ-plot for the X-linked meta-analysis of the African cohorts, including the Gambian, Ghanaian and SA data using the X chromosome specific XWAS software.

**Figure S6:**
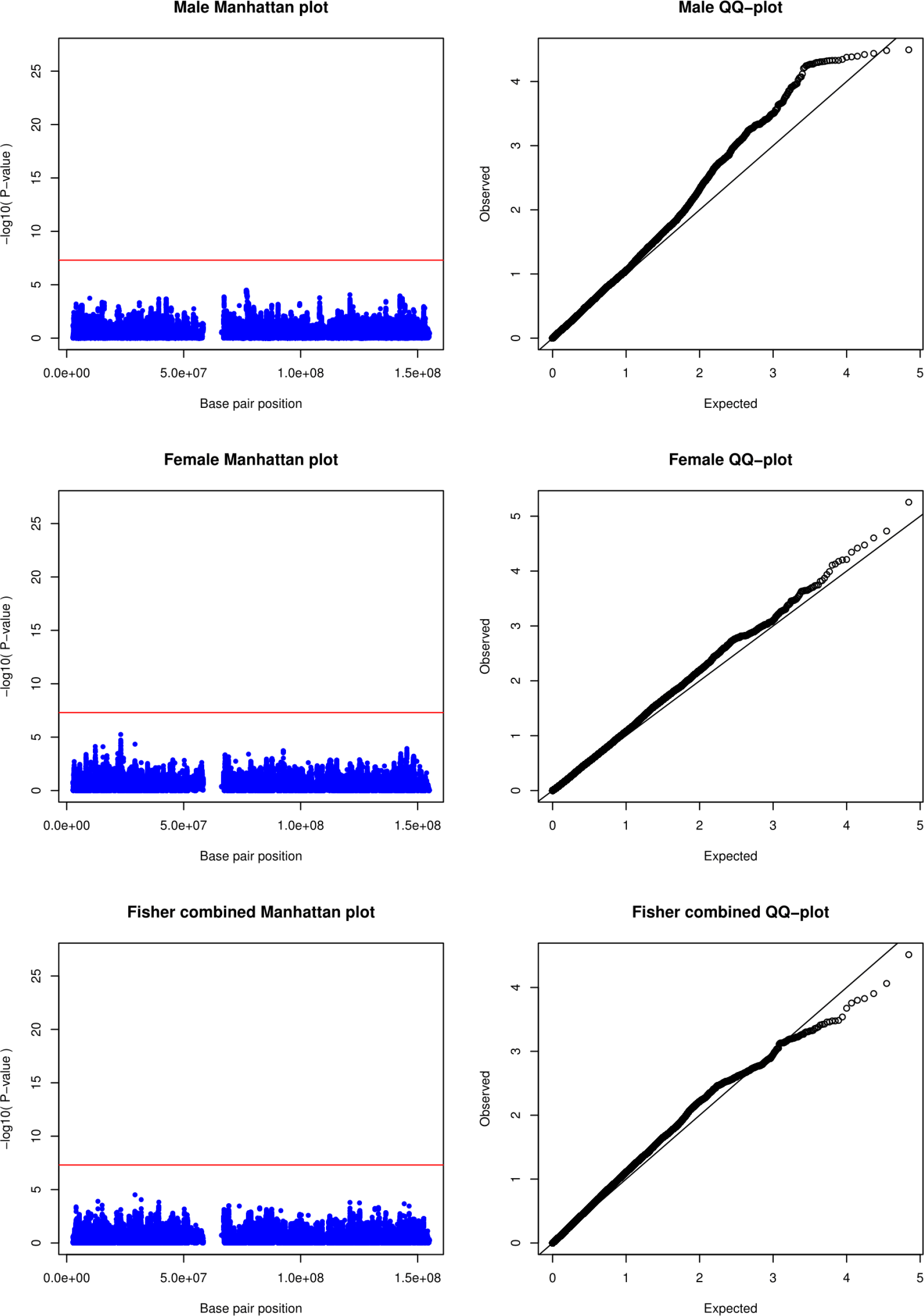
Manhattan and QQ-plot for the X-linked meta-analysis including all cohorts using the X chromosome specific XWAS software.

**Figure S7:**
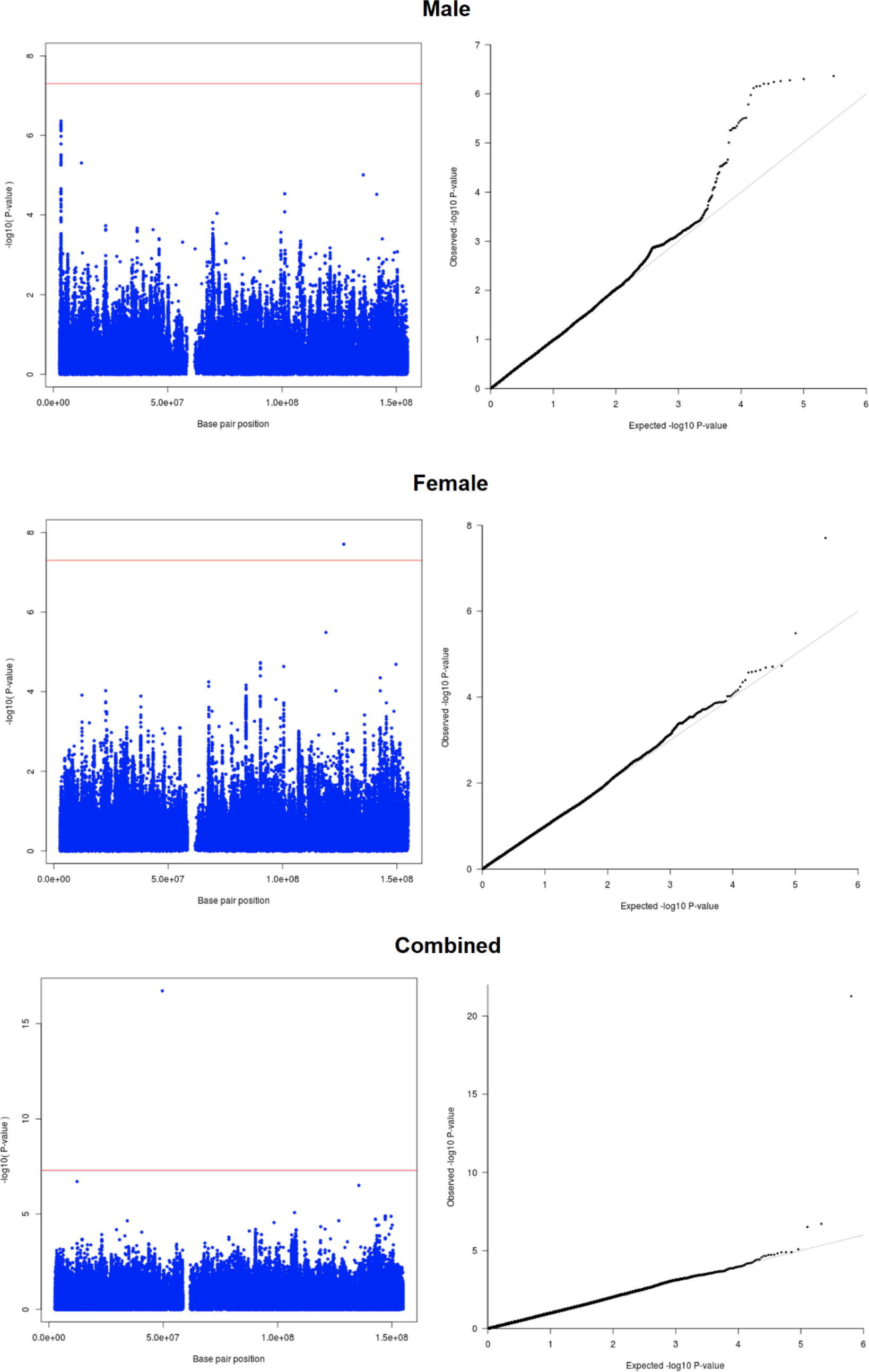
Manhattan and QQ-plot for the X-linked meta-analysis including all cohorts and adjusting for genetic ancestry using the MR-MEGA software.

**Table S1:**
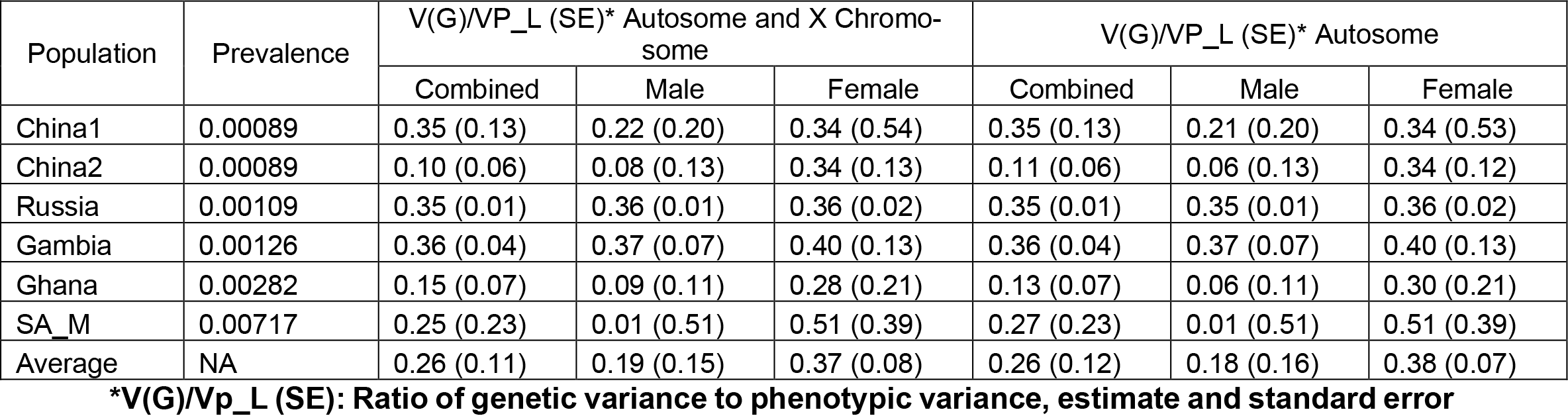
Polygenic heritability estimates for the combined and sex-stratified analysis of all data sets.

**Table S2:**
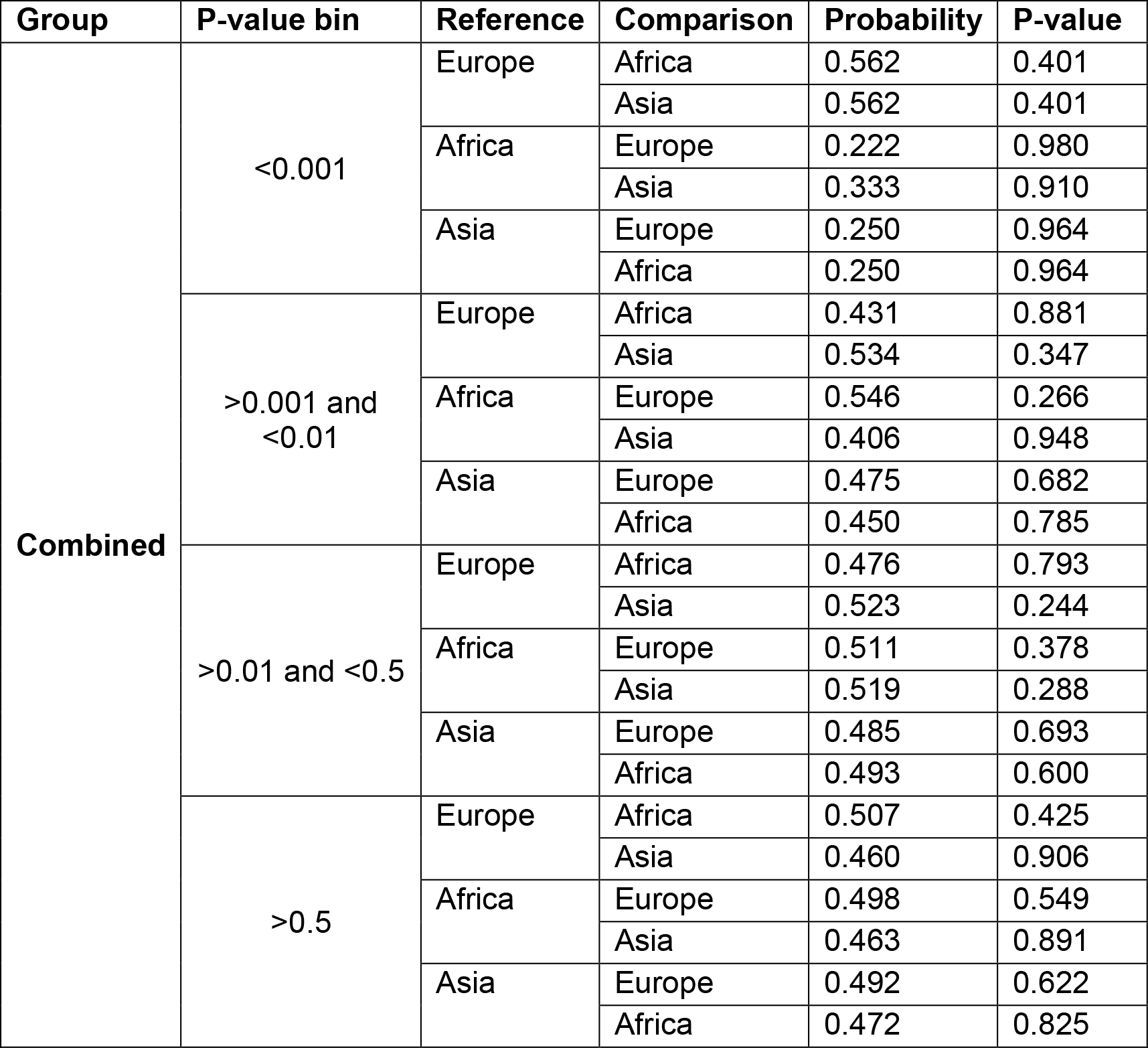
Concordance in direction of effects analysis for the combined data sets.

**Table S3:** Sex-stratified concordance in direction of effects analysis for males.

| Group | P-value bin | Reference | Comparison | Probability | P-value |
| --- | --- | --- | --- | --- | --- |
| **Male** | <0.001 | Europe | Africa | 0.555 | 0.500 |
|  |  |  | Asia | 0.666 | 0.150 |
|  |  | Africa | Europe | 0.571 | 0.500 |
|  |  |  | Asia | 0.428 | 0.773 |
|  |  | Asia | Europe | 0.571 | 0.500 |
|  |  |  | Africa | 1.000 | **0.007** |
|  | >0.001 and <0.01 | Europe | Africa | 0.648 | **0.049** |
|  |  |  | Asia | 0.574 | 0.170 |
|  |  | Africa | Europe | 0.400 | 0.968 |
|  |  |  | Asia | 0.506 | 0.500 |
|  |  | Asia | Europe | 0.530 | 0.306 |
|  |  |  | Africa | 0.510 | 0.459 |
|  | >0.01 and <0.5 | Europe | Africa | 0.462 | 0.863 |
|  |  |  | Asia | 0.476 | 0.790 |
|  |  | Africa | Europe | 0.526 | 0.211 |
|  |  |  | Asia | 0.538 | 0.120 |
|  |  | Asia | Europe | 0.486 | 0.691 |
|  |  |  | Africa | 0.490 | 0.645 |
|  | >0.5 | Europe | Africa | 0.512 | 0.407 |
|  |  |  | Asia | 0.509 | 0.400 |
|  |  | Africa | Europe | 0.496 | 0.573 |
|  |  |  | Asia | 0.536 | 0.132 |
|  |  | Asia | Europe | 0.526 | 0.220 |
|  |  |  | Africa | 0.485 | 0.696 |

**Table S4:** Sex-stratified concordance in direction of effects analysis for females.

| Group | P-value bin | Reference | Comparison | Probability | P-value |
| --- | --- | --- | --- | --- | --- |
| Female | <0.001 | Europe | Africa | 0.454 | 0.725 |
|  |  |  | Asia | 0.272 | 0.967 |
|  |  | Africa | Europe | 0.375 | 0.924 |
|  |  |  | Asia | 0.416 | 0.846 |
|  |  | Asia | Europe | 0.523 | 0.500 |
|  |  |  | Africa | 0.142 | 0.999 |
|  | >0.001 and <0.01 | Europe | Africa | 0.431 | 0.881 |
|  |  |  | Asia | 0.396 | 0.956 |
|  |  | Africa | Europe | 0.494 | 0.583 |
|  |  |  | Asia | 0.545 | 0.227 |
|  |  | Asia | Europe | 0.548 | 0.262 |
|  |  |  | Africa | 0.532 | 0.351 |
|  | >0.01 and <0.5 | Europe | Africa | 0.531 | 0.175 |
|  |  |  | Asia | 0.511 | 0.377 |
|  |  | Africa | Europe | 0.449 | 0.957 |
|  |  |  | Asia | 0.513 | 0.356 |
|  |  | Asia | Europe | 0.521 | 0.267 |
|  |  |  | Africa | 0.463 | 0.893 |
|  | >0.5 | Europe | Africa | 0.490 | 0.647 |
|  |  |  | Asia | 0.540 | 0.114 |
|  |  | Africa | Europe | 0.503 | 0.475 |
|  |  |  | Asia | 0.511 | 0.378 |
|  |  | Asia | Europe | 0.526 | 0.222 |
|  |  |  | Africa | 0.485 | 0.694 |

## Notes

### Competing Interest Statement

The authors have declared no competing interest.

### Author Declarations

A research collaboration agreement was signed by all contributors. Ethics approval for the meta-analysis presented here was granted by the Health Research Ethics Committee of Stellenbosch University (project registration number S17/01/013). In addition, all institutions involved in the ITHGC have ethics approval for their respective studies: China 1 and 2: The study protocol was approved by the Ethics Committee of the Beijing Chest Hospital, the 309 Hospital of the PLA, Shijiazhuang Fifth Hospital, the China PLA General Hospital, the Tongliao TB institute and the Center for Diseases Control and Prevention in Jalainuoer. Russia: Blood samples from all participants were collected and studied with written informed consent according to the Declaration of Helsinki and with approvals from the local ethics committees in Russia (St. Petersburg and Samara) and the UK (Human Biological Resource Ethics Committee of the University of Cambridge and the National Research Ethics Service, Cambridgeshire 1 REC, 10/H0304/71). Gambia: Ethics approval was granted by the Medical Research Council (MRC) and the Gambian government joint ethical committee. Ghana: Ethics approval was granted by the Committee on Human Research, Publications and Ethics, School of Medical Sciences, Kwame Nkrumah University of Science and Technology, Kumasi, Ghana, and the Ethics Committee of the Ghana Health Service, Accra, Ghana. SA A and SA M: Ethics approval was granted by the Health Research Ethics Committee of Stellenbosch University (project registration numbers S17/01/013, NO6/07/132 and 95/072).

